# VirPool: Model-Based Estimation of SARS-CoV-2 Variant Proportions in Wastewater Samples

**DOI:** 10.1101/2022.06.21.22276717

**Authors:** Askar Gafurov, Andrej Baláž, Fabian Amman, Kristína Boršová, Viktória Čabanová, Boris Klempa, Andreas Bergthaler, Tomáš Vinař, Broňa Brejová

## Abstract

**Background:** The genomes of SARS-CoV-2 are classified into variants, some of which are monitored as variants of concern (e.g. the delta variant B.1.617.2 or omicron variant B.1.1.529). Proportions of these variants in a population are typically estimated by large-scale sequencing of individual patient samples. Sequencing a mixture of SARS-CoV-2 RNA molecules from wastewater provides a cost-effective alternative, but requires methods for estimating variant proportions in a mixed sample.

**Results:** We propose a new method based on a probabilistic model of sequencing reads, capturing sequence diversity present within individual variants, as well as sequencing errors. The algorithm is implemented in an open source Python program called VirPool. We evaluated the accuracy of VirPool on several simulated and real sequencing data sets from both Illumina and nanopore sequencing platforms, including wastewater samples from Austria and France monitoring the onset of alpha and delta variants.

**Conclusions:** VirPool is a versatile tool for wastewater and other mixed-sample analysis that can handle both short- and long-read sequencing data. Our approach does not require pre-selection of characteristic mutations for variant profiles, it is able to use the entire length of reads instead of just the most informative positions, and can also capture haplotype dependencies within a single read.

**Availability:** VirPool is an open source software available at https://github.com/fmfi-compbio/virpool.

## Introduction

The pandemic of COVID-19 is accompanied by an unprecedented level of genomic surveillance of the severe acute respiratory syndrome coronavirus 2 (SARS-CoV-2), with more than 10 million genomic sequences deposited in the GISAID database (Elbe and Buckland-Merrett, 2017) by early April 2022. These sequences are mostly obtained from single-patient samples following a positive clinical test. Long-term studies have shown that the composition of viral RNA fragments in wastewater reflects qualitatively and quantitatively the breakdown of virus lineages circulating in the population of the catchment (Agrawal et al, 2022; Amman et al, 2022). Wastewater-based epidemiology (WBE) has recently emerged as a cost effective and scalable alternative to sequencing individual sample patients (Safford et al, 2022; Hrudey and Conant, 2022). Various PCR-based techniques can be used to estimate concentration of virus particles in wastewater to ascertain epidemiological trends. This can be accompanied by sequencing the mixture of virus particles present in the sample, effectively mapping the sequence variation of the virus within a local population. Wastewater monitoring can help to address biases in analysis of clinical samples that are due to differences in test availability and willingness to undergo a clinical test. Moreover, increases in wastewater viral RNA levels can precede the results of clinical testing by several days or even longer when clinical testing is not readily available (Bibby et al, 2021; Hrudey and Conant, 2022), and thus wastewater analysis can provide early warning signals of worsening epidemic situation or emergence of new variants of the virus in a particular area.

Our goal is to identify and quantify the occurrence of variants of SARS-CoV-2 within pooled samples (which also include wastewater samples). In our work, we rely on Pango lineage classification of virus variants (Rambaut et al, 2020), which is based on phylogenetic analysis of sequenced genomes. Selected clades of the phylogenetic tree are assigned identifiers, leading to a hierarchical system of subclades nested in larger clades. The World Health Organization monitors prevalence of variants around the world and selects variants of concern (VOCs) characterized by an increased transmissibility or virulence or the ability to evade protection provided by vaccines and drugs. Several of these variants have caused massive epidemic waves, most notably the Alpha, Delta, and Omicron variants (Pango lineages B.1.1.7, B.1.617.2, and B.1.1.529, respectively). It is therefore of high interest to monitor the prevalence of these variants, particularly at an onset of a new wave, when the public health authorities expect the arrival of a new variant in a certain area.

Early work in analysis of SARS-CoV-2 wastewater samples concentrated on producing an overall consensus sequence of the sample or on detection of individual mutations followed by manual analysis of the results (Xie et al, 2022; Izquierdo-Lara et al, 2021; Crits-Christoph et al, 2021; Nemudryi et al, 2020; Hillary et al, 2021; Agrawal et al, 2021, 2022). Later, the presence of variants of concern was detected based on pre-selected mutations typical for each variant (La Rosa et al, 2021; Jahn et al, 2021; Fontenele et al, 2021).

In order to also quantify the prevalence of a particular variant, the most straightforward approach uses again several pre-selected sites with mutations characteristic for the variant. At each such site, the proportion of the allele belonging to the variant is estimated, and the final proportion is determined as a mean or a median of single-site estimates (Wurtz et al, 2021; Rios et al, 2021b; Brunner et al, 2022; Pechlivanis et al, 2022). Since each variant is considered independently of others, the method can produce inconsistent estimates (e.g. the sum of proportions of individual variants is greater than one) and can be biased by mutations shared by multiple variants, an issue which is likely to be exacerbated given the increased occurrence of convergent evolution events between different lineages due to selective pressures. To account for these shared mutations, Ellmen et al (2021) estimate the proportions of variants by optimising the L2 metric between a mixture of base frequencies of individual variants and observed frequencies of specific mutation sites. Amman et al (2022) pushed this idea further by estimating the proportions jointly for multiple samples, taking the time of their collection into account.

A similar problem has been previously addressed in the context of virus populations. In the quasispecies spectrum reconstruction (QSR) problem, the aim is to analyze a sequencing sample containing reads from several distinct virus variants (also called haplotypes) to recover individual haplotype sequences and quantify their prevalence (Eriksson et al, 2008; Zagordi et al, 2011; Ahn and Vikalo, 2018). Typically, haplotypes are recovered by specialized assembly algorithms employing read overlaps and sequence coverage (see e.g. the path cover approach in ShoRAH (Zagordi et al, 2011)), followed by quantification of individual variants. In general, these approaches assume that individual haplotypes yield a consistent coverage across the whole reference genome, that the sequencing reads are randomly sampled from the haplotypes, and that haplotypes themselves are well represented by a single consensus sequence, possibly with a few local variations attributable to sequencing errors.

However, these assumptions are not satisfied in the case of SARS-CoV-2 wastewater sequencing. SARS-CoV-2 wastewater samples are typically sequenced by ARTIC protocol, originally developed in the context of Zika virus epidemics (Quick et al, 2017). The virus sequence is divided into overlapping segments (called *amplicons*) that are first amplified through PCR, so as to increase the number of molecules present in the sequencing sample. Depending on the protocol, the approximate length of amplicons varies between 400bp (Loman et al, 2020) (further referred to as *short amplicons*) and 1.5-2.5kbp (Resende et al, 2020; Eden and Sim, 2020; Freed et al, 2020) (*long amplicons*). Only after the PCR amplification, the pooled sample is sequenced by Illumina or Oxford Nanopore MinION sequencers. The efficiency of the PCR amplification varies between amplicons, resulting in highly uneven coverage along the genome. Moreover, sequencing reads do not span amplicon boundaries, breaking the haplotype linkage between the amplicons. Finally, individual SARS-CoV-2 variants (as defined by Pango lineages) typically include a large number of distinct sequences, and this diversity is difficult to express as a single consensus sequence.

In this paper, we introduce a new approach based on a probabilistic model of sequencing reads originating from a mixture of variants. Our model captures sequence diversity present within individual variants through employing *variant profiles* derived from available GISAID sequences for a particular variant. At the same time, we also model sequencing errors, which is essential in application to data sets obtained by sequencing technologies with higher error rates, such as nanopore sequencing. Our approach does not classify individual reads or sites as belonging to a particular variant, but instead searches for a solution that has the highest consistency with the observed data. Consequently, we do not require pre-selection of sites characteristic for each variant, and we can use the information contained in the full length of the sequenced reads. In this aspect, our approach is similar to the approach by Eriksson et al (2008), though our model is more complex due to the specifics of the wastewater analysis problem. Finally, our approach is able to exploit linkage between individual sites within the same sequencing read, which leads to an increased accuracy in case of using long nanopore amplicons in spite of higher sequencing error rates of nanopore sequencing. An early version of our model (Gafurov et al, 2021), which was unable to take this information into account, suffered from lower accuracy which had to be addressed by ad-hoc heuristics. We have tested our software on analysis of both simulated and real data sets and we have shown that our approach outperforms the median approach previously employed for estimating proportions of SARS-CoV-2 variants in wastewater samples.

## Results

### Mixture model for variant proportion estimation

For a given sequencing sample, VirPool estimates the fraction of sequencing reads originating from selected variants. Variant *k* is characterized by its *variant profile P*_*k*_, where *P*_*k*_(*i, a*) is the probability of observing nucleotide *a* at position *i* in variant *k*. The positions are numbered according to the reference genome, and all sequences that we consider are aligned to this reference genome.

The VirPool algorithm is governed by a probabilistic model that assigns a likelihood to weights of individual variants *w*_1_, …, *w*_*K*_, where *K* is the number of variant profiles. Intuitively, these weights correspond to proportions of sequencing reads originating from individual variants mixed in a particular sample. Assuming that a particular read *r*, starting at position *s*, originates from a variant *k*, the probability of observing this read is simply

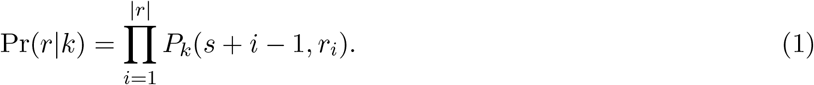

This probability needs to be also adjusted for sequencing errors (for details, see Methods).

The overall likelihood is the probability of observing sequence of reads *R* = (*r*_1_, …, *r*_*m*_) given the variant weights *w*_1_, …, *w*_*k*_. In our model, each read is generated independently; thus the likelihood can be expressed as

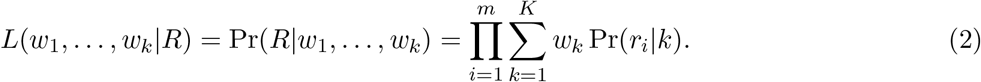

Here we assume that starting positions and lengths of individual reads are fixed in advance. Note that if these were sampled from any distribution independent of variants, this would only add a constant factor to the likelihood and would have no influence on the weight optimization.

For a given set of sequencing reads and variant profiles, the weights are estimated so that the likelihood *L*(*w*_1_, …, *w*_*k*_|*R*) is maximized. For details of the optimization algorithm, see Methods.

The results of VirPool analysis are dependent on the selection of variants included in the analysis.

Typically, one would select variants of interest circulating in a given region at a considered time. In our experiments, we also include variant “other” that represents all the remaining SARS-CoV-2 sequences not belonging to the selected variants. High weight of the “other” variant in the result allows the user to detect that the set of the variants should be adjusted.

### Accurate prediction of variant proportions for different sequencing technologies

To evaluate the accuracy of our methods, we have prepared synthetic mixtures combining several single-patient sequencing read sets downloaded from public databases, each containing sequencing reads from a single virus variant. We have selected four variants common in Europe in the Fall of 2020 (B.1.1.7, B.1.160, B.1.177, B.1.258) as well as samples from other variants which should be correctly classified as “other” profile (B.1.221, B.1.1.170, B.1.367, B.1.1.37, AP.1); see Methods for the mixture creation details. We have estimated proportions of these profiles both by VirPool and by the baseline median method described in Methods and compared the results to true proportions.

Table 1 shows that in almost all cases, VirPool can accurately estimate the true proportions. This is true for all three considered sequencing protocols (Illumina paired 150bp reads with 400bp amplicons, Oxford nanopore short 400bp amplicons, and Oxford nanopore long 2kbp amplicons). The method also worked well when the mixtures contained variants included in the “other” combined profile. The median method (Rios et al, 2021b), while providing in many cases similar results to VirPool, systematically overestimates the explicitly listed variants, in many cases resulting in estimates of named variants summing to more than 1.

**Table 1:** Comparison of true and estimated variant proportions for the synthetic mixtures. The first group corresponds to mixtures created with Oxford Nanopore reads with long (2 kbp) amplicons, the second group corresponds to mixtures created with Oxford Nanopore reads with short (400 bp) amplicons, and the third group corresponds to mixtures created with Illumina reads. The proportion of other for the median estimator is calculated as 1 minus the sum of the estimated proportions (therefore it can be negative).

| mixture name |  | B.1.1.7 | B.1.160 | B.1.177 | B.1.258 | other |
| --- | --- | --- | --- | --- | --- | --- |
| ont-long-1 | true | 0.19 | 0.42 | 0.23 | 0.16 | 0.00 |
|  | VirPool | 0.18 | 0.42 | 0.23 | 0.17 | 0.00 |
|  | median | 0.19 | 0.43 | 0.24 | 0.17 | -0.03 |
| ont-long-2 | true | 0.17 | 0.38 | 0.21 | 0.15 | 0.09 |
|  | VirPool | 0.16 | 0.38 | 0.22 | 0.15 | 0.09 |
|  | median | 0.17 | 0.38 | 0.20 | 0.15 | 0.09 |
| ont-long-3 | true | 0.42 | 0.00 | 0.00 | 0.35 | 0.23 |
|  | VirPool | 0.41 | 0.00 | 0.00 | 0.37 | 0.22 |
|  | median | 0.39 | 0.01 | 0.02 | 0.36 | 0.23 |
| ont-long-4 | true | 0.00 | 0.65 | 0.35 | 0.00 | 0.00 |
|  | VirPool | 0.00 | 0.64 | 0.36 | 0.00 | 0.00 |
|  | median | 0.01 | 0.63 | 0.43 | 0.02 | -0.08 |
| ont-short-1 | true | 0.19 | 0.42 | 0.23 | 0.16 | 0.00 |
|  | VirPool | 0.18 | 0.42 | 0.23 | 0.17 | 0.00 |
|  | median | 0.19 | 0.43 | 0.24 | 0.17 | -0.03 |
| ont-short-2 | true | 0.17 | 0.38 | 0.21 | 0.15 | 0.09 |
|  | VirPool | 0.16 | 0.38 | 0.22 | 0.15 | 0.09 |
|  | median | 0.17 | 0.38 | 0.20 | 0.15 | 0.09 |
| ont-short-3 | true | 0.42 | 0.00 | 0.00 | 0.35 | 0.23 |
|  | VirPool | 0.41 | 0.00 | 0.00 | 0.37 | 0.22 |
|  | median | 0.39 | 0.01 | 0.02 | 0.36 | 0.23 |
| ont-short-4 | true | 0.00 | 0.65 | 0.35 | 0.00 | 0.00 |
|  | VirPool | 0.00 | 0.64 | 0.36 | 0.00 | 0.00 |
|  | median | 0.01 | 0.63 | 0.43 | 0.02 | -0.08 |
| illumina-1 | true | 0.15 | 0.23 | 0.27 | 0.34 | 0.00 |
|  | VirPool | 0.16 | 0.26 | 0.29 | 0.29 | 0.00 |
|  | median | 0.20 | 0.26 | 0.30 | 0.35 | -0.11 |
| illumina-2 | true | 0.13 | 0.20 | 0.23 | 0.29 | 0.16 |
|  | VirPool | 0.14 | 0.22 | 0.25 | 0.25 | 0.13 |
|  | median | 0.17 | 0.23 | 0.25 | 0.30 | 0.06 |
| illumina-3 | true | 0.16 | 0.25 | 0.00 | 0.37 | 0.21 |
|  | VirPool | 0.18 | 0.29 | 0.01 | 0.33 | 0.19 |
|  | median | 0.23 | 0.32 | 0.00 | 0.38 | 0.07 |
| illumina-4 | true | 0.00 | 0.46 | 0.54 | 0.00 | 0.00 |
|  | VirPool | 0.00 | 0.47 | 0.53 | 0.00 | 0.00 |
|  | median | 0.00 | 0.49 | 0.66 | 0.00 | -0.16 |

### Prediction accuracy at low coverages

To evaluate the accuracy at lower coverages, we have subsampled our synthetic mixtures. Figure 1 shows the mean squared error of predictions averaged over multiple subsamples. Even though nanopore sequencing has a much higher sequencing error rate than Illumina, the best weight estimates are achieved with nanopore long 2kb amplicons. This suggests that VirPool can effectively take advantage of long-range dependencies between positions covered by a single amplicon. For both nanopore and Illumina with short amplicons, the error decreases with increasing coverage until reaching a plateau, in most cases around 100× genome coverage. In most cases, the median approach has significantly larger error than VirPool.

**Figure 1:**
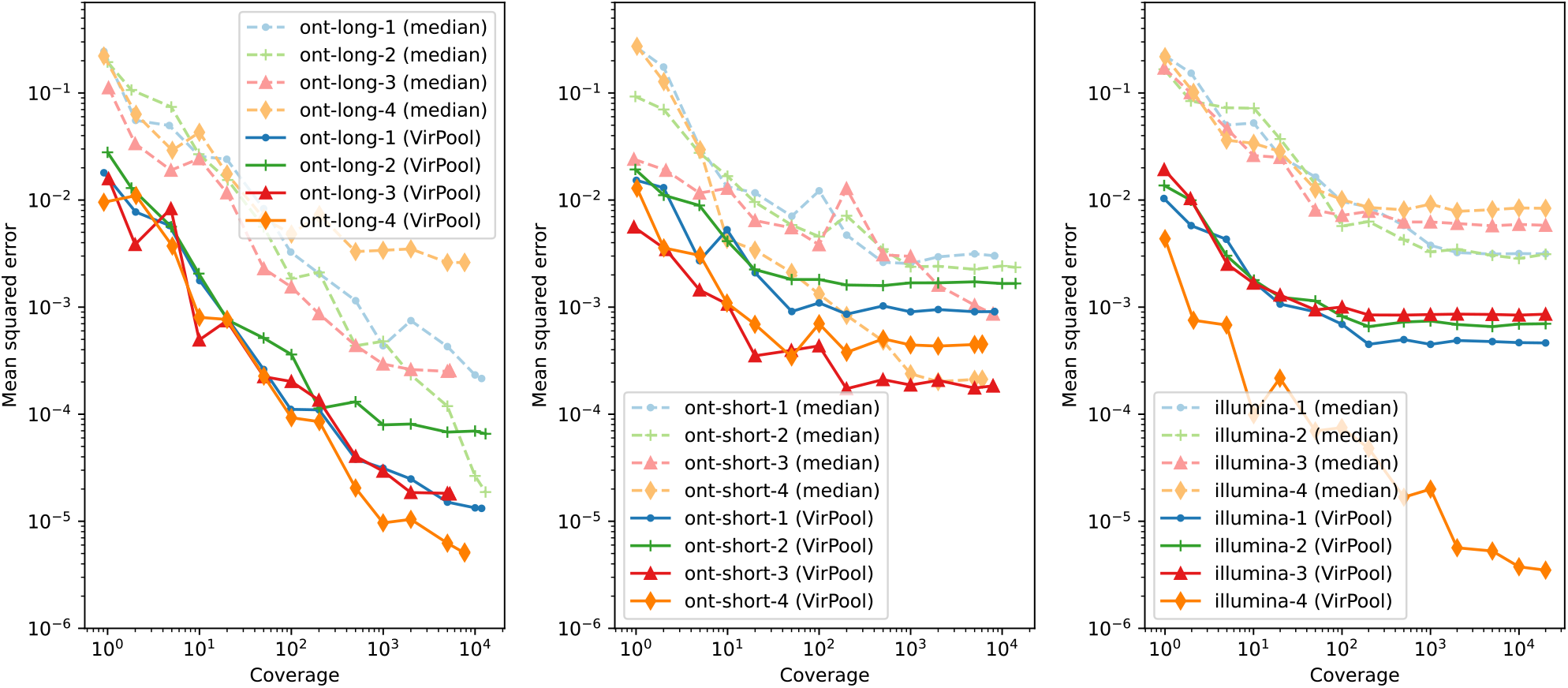
Variant proportion estimation errors as a function of sequencing coverage. Left: long nanopore mixtures; Center: short nanopore mixtures; Right: Illumina mixtures. The mean squared error (MSE) was averaged over ten random subsamples of synthetic mixtures from Table 1 for each considered coverage. Both axes are in logarithmic scale.

### Reliability of detection of low-abundance variants

To test detection of variants occurring at low frequencies, we have created artificial mixtures of two variants from among the Fall of 2020 samples, with the minor variant present in frequencies ranging from 0.1% to 20%. Figure 2 shows that VirPool is generally accurate for frequencies of 5% or more. Even frequencies of 2% were generally detected, but the variance in the results is high. Conversely, at very low frequencies, the presence of the minor variant may be overestimated. Note that detection of such low-frequency variants has been shown to be difficult and requires an extremely high coverage even with Illumina sequencing data (Van Poelvoorde et al, 2021).

**Figure 2:**
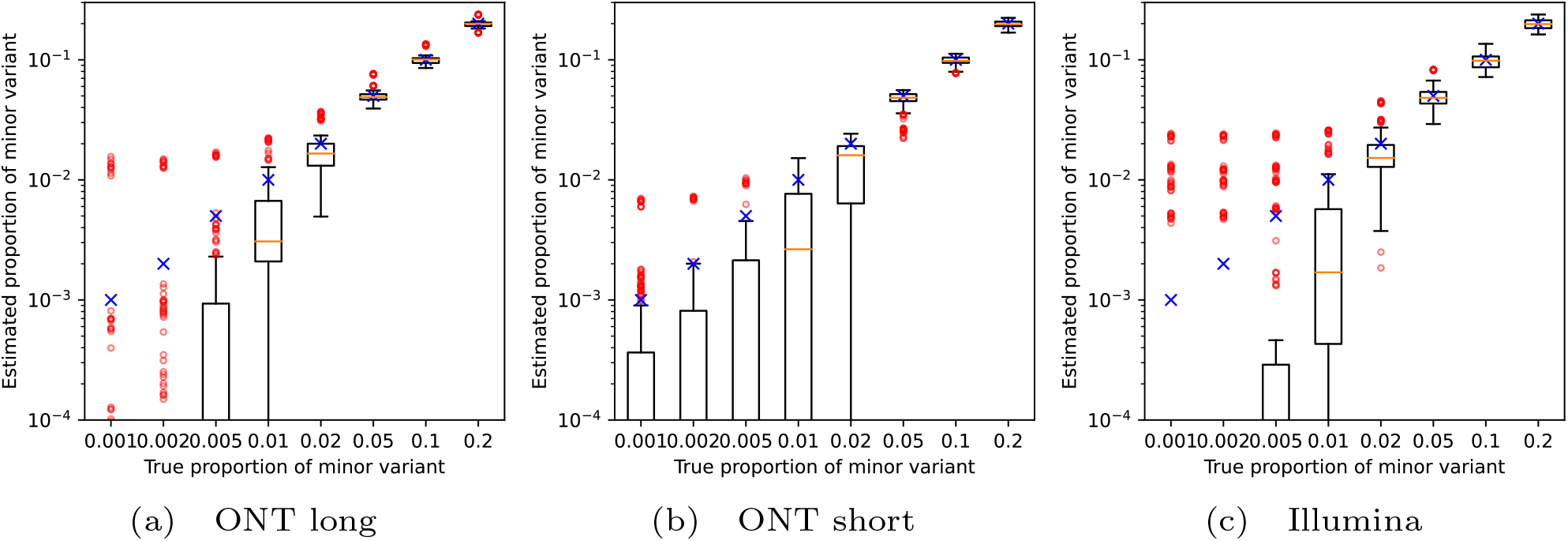
Estimated proportion of the minor variant in synthetic experiments among Fall 2020 samples. Blue crosses represent the true proportion of the minor variant. Orange lines represent the median, red circles are outliers. In each setting, the graphs show the distribution for 10 randomly generated data sets for each pair of variants (including the “other” group). The average coverage for each synthetic data set was 5000.

We have also attempted to replicate data corresponding to the Alpha (B.1.1.7) wave transitioning to Delta (B.1.617.2) (Figure 3) and Delta transitioning to Omicron (B.1.1.529, specifically BA.1 subvariant) (Figure 4). Again, results above 5% are reliable, with the exception of short nanopore data in the Omicron wave, where the estimated weights of Omicron are lower due to a false prediction of the “other” variant.

**Figure 3:**
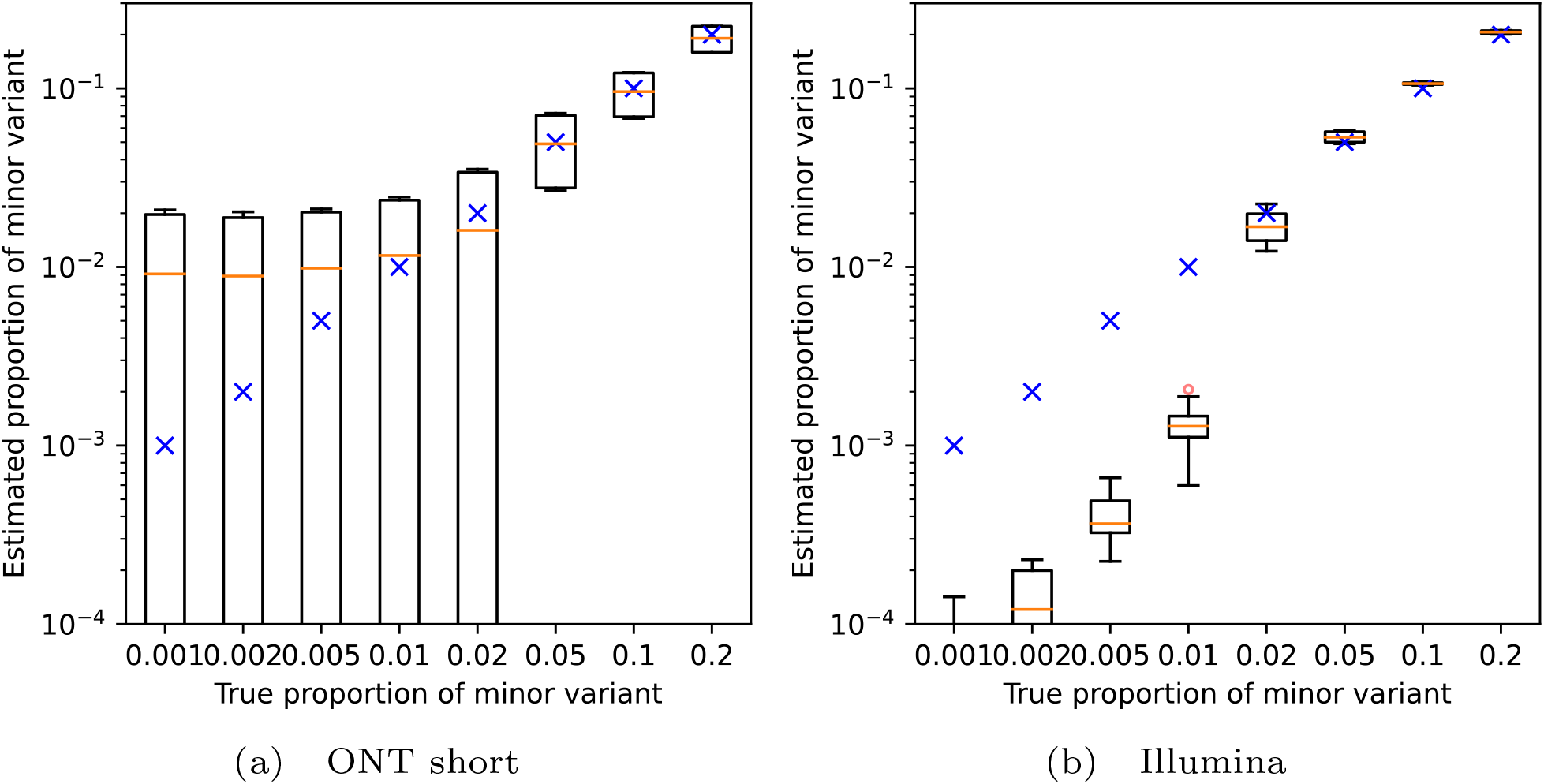
Estimated proportion of the minor variant in synthetic mixtures of the Alpha and Delta variants. Blue crosses represent the true proportion of the minor variant. Orange lines represent the median, red circles are outliers. In each setting, the graphs show the distribution for 20 randomly generated data sets with the prescribed minor variant proportion, with Alpha and Delta playing the role of the minor variant in 10 samples each. The average coverage for each synthetic data set was 5000.

**Figure 4:**
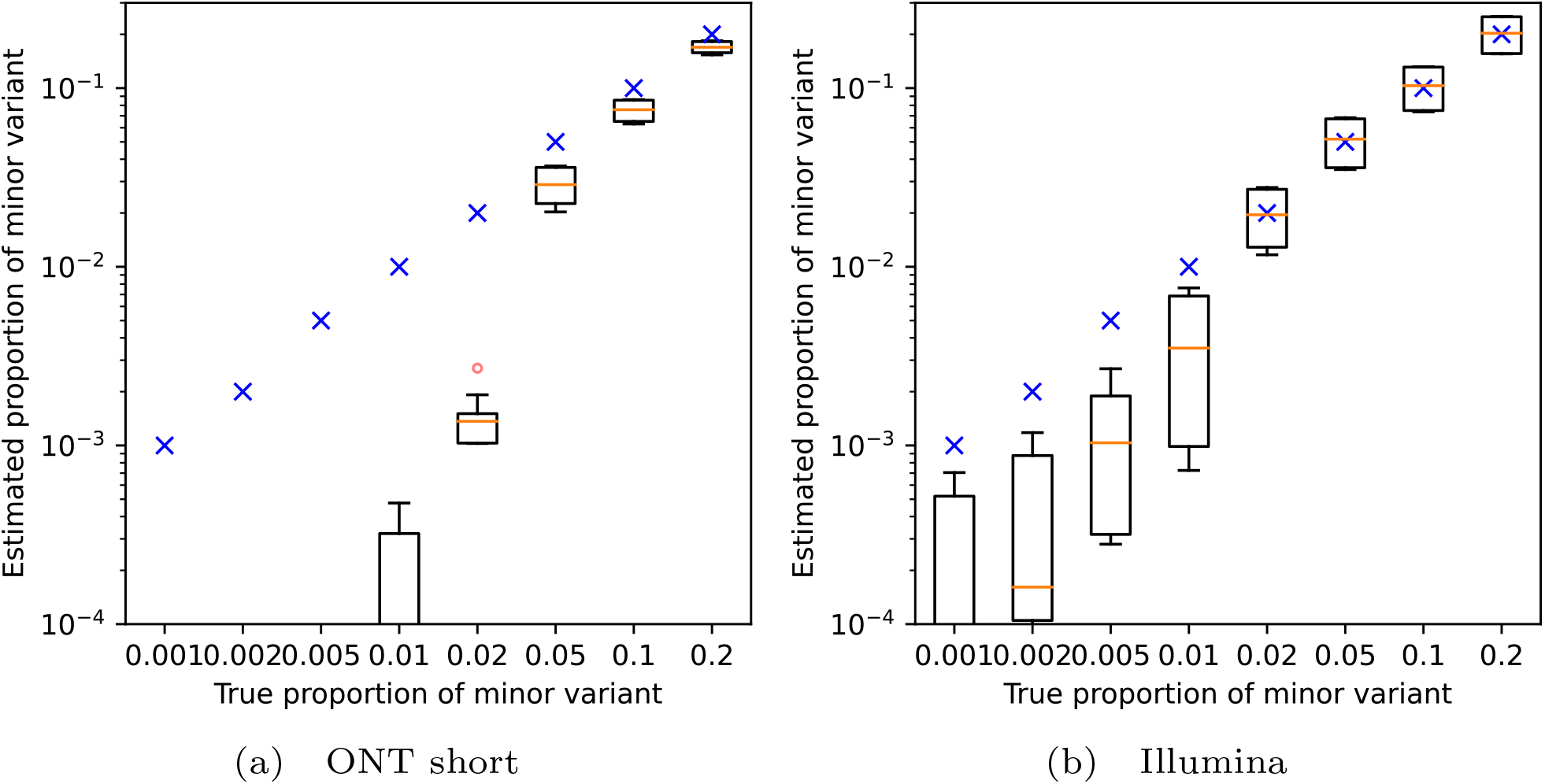
Estimated proportion of the minor variant in synthetic mixtures of the Delta and Omicron variants. Blue crosses represent the true proportion of the minor variant. Orange lines represent the median, red circles are outliers. In each setting, the graphs show the distribution for 20 randomly generated data sets with the prescribed minor variant proportion, with Delta and Omicron playing the role of the minor variant in 10 samples each. The average coverage for each synthetic data set was 5000.

### In-vitro mixture of patient samples

Using nanopore long amplicons, we have sequenced and analyzed a mixture of eight clinical samples. Presence or absence of individual variants was correctly identified by VirPool (Table 2), and the estimated proportions agree well with a separate analysis based on examination of frequencies of mutations specific for individual clinical samples, as determined by previous sequencing of each sample individually. Note that, even though an effort has been made to balance the original sample proportions using dilution factors based on measured Cq values (Supplementary Table S2), the resulting proportions are influenced by many factors, such as different levels of fragmentation of RNA and subsequent differences in amplification efficiency.

**Table 2:** Estimated proportions for the in-vitro mixture sample of eight patient samples. The second rows shows a simple estimate based on computing the median of frequencies for mutations unique to one of the eight samples and then summing the medians over the samples with the same variant.

|  | B.1.1.7 | B.1.160 | B.1.177 | B.1.258 | other |
| --- | --- | --- | --- | --- | --- |
| number of samples | 2 samples | 1 sample | 0 samples | 4 samples | 1 sample |
| estimate from unique mutations | 0.55 | 0.08 | 0.00 | 0.33 | 0.07 |
| VirPool estimate | 0.53 | 0.07 | 0.00 | 0.25 | 0.15 |

### Analysis of wastewater samples from the Alpha wave

We have applied VirPool to time series data sets of wastewater samples spanning several months in regions of Bischofshofen, Austria (Illumina data) (Amman et al, 2022) and Nice, France (nanopore data) (Rios et al, 2021b). Wastewater samples are highly challenging, because the coverage is often highly uneven (see Supplementary Figure S1).

Figure 5 shows the analysis of a time series sampled between December 2020 and February 2021 from the area of Bischofshofen, Austria (State of Salzburg), sequenced by Illumina short read protocol. Comparing VirPool analysis to the analysis by VaQuERo pipeline (Amman et al, 2022), both tools predict a sharp increase in the Alpha variant in January and February of 2021, which also apparent in the clinical samples from GISAID.

**Figure 5:**
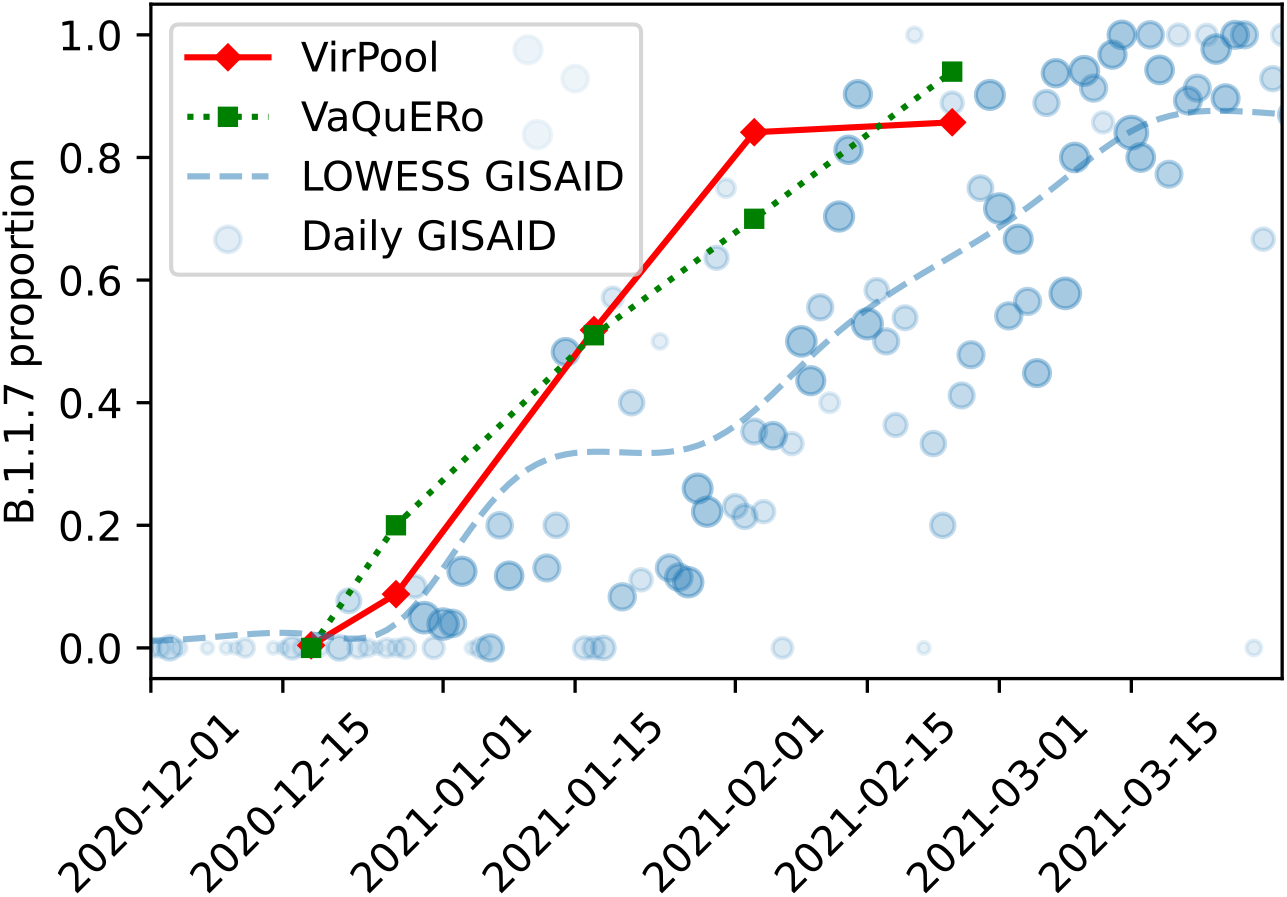
Estimated proportions of the Alpha (B.1.1.7) variant in wastewater samples from Bischofshofen, Austria (Amman et al, 2022). The red line represents the proportions estimated by VirPool model. The green line represents the proportions estimated by VaQuERo model (Amman et al, 2022). The blue circles represent the daily proportions of B.1.1.7 among Austrian samples submitted to GISAID, the number of samples is reflected by circle size; blue line is the smoothed version of those data.

Both methods also agree on the general composition of samples and most of the trends (Supplementary Table S4). One of the notable differences is a lower prevalence of the Alpha variant at the end of February in VirPool predictions, which is compensated by a rebound in the B.1.258 prevalence to 10%. By examining allele frequencies, we have found 7 alleles that strongly support inclusion of B.1.258 (Supplementary Table S5); three of these alleles are typical for the whole B.1.258 clade, while four additional alleles are characteristic for subvariant B.1.258.17 that has been observed in clinical samples in the state of Salzburg (12 samples out of 136 in GISAID in February and March). In contrast, several alleles very common in B.1.258 are either missing completely (e.g., 8047T) or are present at very low frequencies (e.g., 7767C, 22879A, 29734C). This may be a consequence of a very uneven coverage or variant-specific differences in amplification efficiency for specific primers, may possibly indicate recombination, or it may indicate some other variant sharing characteristic mutations with B.1.258.17; ten samples in GISAID outside of B.1.258 collected between January and March share the same 7 alleles. Their classification is either generic (B.1), some of them are classified as B.1.367 or B.1.221. In spite of this uncertainty, the data seem to support a lower prevalence of the Alpha variant and possible presence of B.1.258.17 variant in agreement with the VirPool predictions.

One of the strengths of VirPool is that the same method can be applied to both Illumina and nanopore data sets, only changing settings for the sequencing error rate. Figure 6 shows the result of analysis of selected samples from Nice, France, sequenced by the short-amplicon nanopore sequencing protocol. In agreement with Rios et al (2021b), we observe a very sharp increase in the Alpha variant prevalence in February 2021, which is not observed in the clinical samples from GISAID in the Provence-Alpes-Côte d’Azur region, suggesting that this outbreak was not captured in a timely manner by genome sequencing efforts. In agreement with Rios et al (2021b) we see a significant prevalence of the Alpha variant in the Les Moulins site already in January 2021.

**Figure 6:**
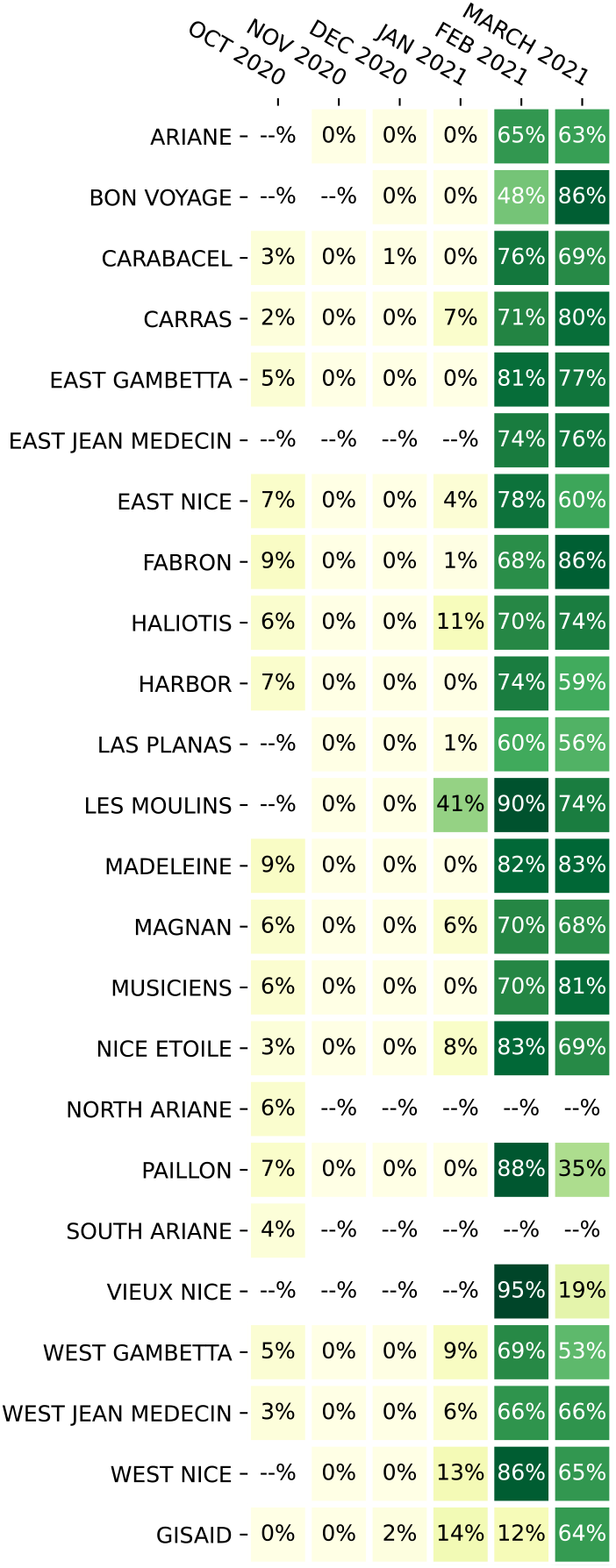
Estimated the Alpha (B.1.1.7) proportions in wastewater samples from Nice, France, using data from Rios et al (2021b). If multiple samples were sequenced for a given location and month, the median was taken. The last row (“GISAID”) is the proportion of sequences with variant B.1.1.7 submitted to GISAID database from French region Provence-Alpes-Ĉote d’Azur in the same month.

VirPool consistently predicts several percent of the Alpha variant in all samples from October 2020. Rioset al (2021b) also observe several mutations characteristic for the Alpha variant in October 2020, but do not comment or investigate this phenomenon. Upon closer examination, we see a similar pattern as in the case of B.1.258 variant in the Austrian samples, with some mutations characteristic for a given variant (in this case Alpha) having a relatively high frequency, while others being absent (Supplementary Table S6). Here, having samples from multiple locations, we observe that some genome positions give consistent results, while others vary between samples, further supporting the hypothesis of amplification efficiency differences. Overall, we hypothesize that the Alpha variant was indeed present in the area, which is further supported by three GISAID Alpha samples collected in Marseille in the same month.

VirPool also estimated a significant decrease in the Alpha variant proportions in several locations between February and March 2021. We have investigated this decrease in the Harbor location (from 74% in February to 59% in March), where VirPool estimated a rebound of variants B.1.177 (to 11%) and B.1.160 (to 15%). The predicted increase in B.1.177 seems to be driven by C22227T mutation, which is characteristic for B.1.177 and very rare in B.1.1.7; nonetheless, it appeared at the frequency of 69% in the reads from this sample (Supplementary Table S7). All the other mutations characteristic for B.1.177 are either present at much lower frequencies or completely missing. Also, C22227T mutation occurs in several B.1.1.7 GISAID samples from France. All this evidence leads us to a conclusion that a B.1.1.7 subvariant with this mutation has been highly prevalent in this area and the rebound of B.1.177 is a false positive. This points to a weakness of variant characterization by probabilistic profiles representing global distribution of mutations, which may not agree with locally circulating strains, although as we see in other experiments, this usually does not cause problems in the estimation.

## Methods

### Variant profile estimation

Our method requires a set of *K* profiles, each representing one variant of the SARS-CoV-2 virus. In these profiles, *P*_*k*_(*i, a*) is the probability of observing nucleotide *a* at position *i* in variant *k*, where positions are numbered according to the reference sequence (we use Wuhan/Hu-1/2019).

In our experiments, we build these profiles from the SARS-CoV-2 genomic sequences downloaded from the GISAID database (Elbe and Buckland-Merrett, 2017). We have used GISAID version from February 5, 2022, omitting sequences with incomplete or missing collection date, and incomplete genomes with less than 25kbp of sequence. We have then subsampled the data so that at most roughly 50,000 sequences were kept per month. In specific experiments, we use only a subset of GISAID records corresponding to the period from which our samples originate.

In each experiment, we selected several variants designated by their Pango lineage identifier (Supplementary Table S1). The profile for each selected variant is built from the samples assigned to this lineage and its sublineages according to GISAID metadata. Specifically, we assign each sequence to the nearest ancestor clade from our list of selected lineages. This allows for selecting both a lineage and its sublineage, such as B.1.1 and B.1.1.7; in such case sequences from the sublineage are excluded from the parent lineage profile. If no ancestor clade is in the list, the sequence is assigned label “other”, representing genomic background of all other lineages.

Each sequence was aligned to the common reference sequence Wuhan/Hu-1/2019 by minimap2 (Li, 2018). Intuitively, value *P*_*k*_(*i, a*) would be estimated from data as the relative frequency of symbol *a* at position *i* among the genomes assigned to the variant *k*. However, some variants contain characteristic deletions shared by almost all genomes belonging to the variant. As a result, some genomic positions are covered by only a small number of genomes belonging to variant *k*. Let *γ*_*k*_ be a threshold on the coverage. Then we set the variant profile probabilities as follows:

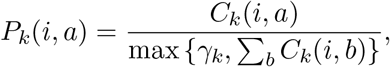

where *C*_*k*_(*i, a*) is the number of occurrences of base *a* at position *i* among genomes of variant *k*. Thus, the positions with coverage lower than *γ*_*k*_, typically containing gaps characteristic for the variant, will have the sum of values *P*_*k*_(*i, a*) lower than one. This will have no impact on reads originating from variant *k*, because they have gaps at such positions, but it will penalize reads from other variants that do not have such gaps.

In our experiments, we set *γ*_*k*_ as the coverage at the first percentile (smallest 1%) of coverage within the genome for each variant *k*.

To select the list of variants for analysis of new samples, the user would choose ones that circulate in a particular area at the time, with additional variants added from relevant watch lists (such as WHO variants of concern and variants of interest). The profiles should be built based on sequences with recent collection dates (latest months) and updated periodically in order to include recent evolutionary changes within virus sequences.

### Substitution error

The mixture model characterized by equations (1) and (2) in the Results section assumed error-free sequencing. In our final model, we add substitution sequencing errors that occur uniformly at random with error rate *ε*. The probability of observing read *r* at position *s* from variant *k* is then

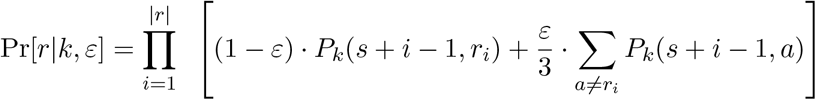

In our experiments, we use *ε* = 0.001 for Illumina reads and *ε* = 0.05 for nanopore reads. Note that both real insertions and insertion errors are ignored by our model; read positions with a deletion are treated as missing data.

### Mixture weight estimation

The optimal weights *W* = *w*_1_, …, *w*_*K*_ are estimated via minimisation of the negative log-likelihood

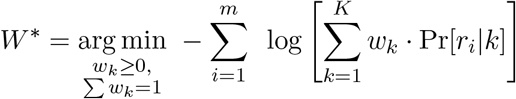

To simplify the optimisation task to an unconstrained case, we use *softmax transformation* (Bridle, 1990). Namely, we define auxiliary variables *ξ*_1_, …, *ξ*_*K*_ and set the values of the original weight variables *w*_1_, …, *w*_*K*_ as the softmax of the auxiliary variables:

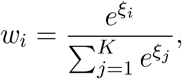

This transformation ensures that all weight variables are non-negative and sum up to 1. This leads to the following unconstrained optimization task:

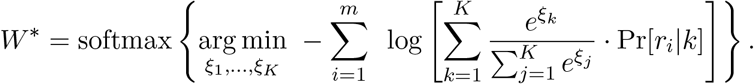

The minimisation is done using the L-BFGS-B algorithm (Zhu et al, 1997) implemented in scipy Python library (Virtanen et al, 2020) with symbolic Jacobian. We use `numba` library to speed up the calculations by pre-compilation of time-critical functions.

### Analysis of sequencing samples

Alignments of sequencing reads to the reference genome were downloaded from ENA, or where needed, were produced by `minimap2` for ONT reads and `bwa-mem` for Illumina reads. We use only primary alignments for each read. In our analysis, we consider a paired read as a single unit with both parts originating in the same variant, and we require that both parts are aligned.

Some mutations tend to occur repeatedly within the virus evolution (homoplasic sites). To avoid confusion between variants, we mask known homoplasic sites (De Maio et al, 2020) and do not consider them in the analysis. Some variants have mutations within primer binding positions. These positions will appear in the reads as matching the primer, not the sequenced sample. Since in most cases, these primers are not trimmed in the underlying datasets, we mask primer positions (ARTIC protocol V3 or V4 as appropriate).

### Baseline method and evaluation

In our experiments, we compare VirPool with an estimator based on median proportions previously used by Rios et al (2021b); similar methods were also used by other studies (Wurtz et al, 2021; Pechlivanis et al, 2022; Brunner et al, 2022). For each selected variant, this estimator needs a list of characteristic mutations; we use the lists by Rios et al (2021b) from their code repository Rios et al (2021a). The relative frequency of each characteristic mutation is computed among reads aligned to the corresponding position. The resulting estimator of the variant’s proportion is the median of these frequencies. Since the median estimate is computed independently for each variant, it is not guaranteed that proportions of all variants will sum to one. We attribute the remaining probability to the background “other” variant; this can also be a negative number.

To evaluate the accuracy of both VirPool and the median estimator on simulated data, we use the *mean squared error* (*MSE*) measure defined as 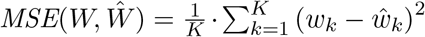,where *W* = (*w*_1_, …, *w*_*K*_) and 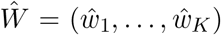 are the true and estimated proportions of variants, respectively. Note that this metric supports negative weight estimates, so it can be used for the median estimator.

### Synthetic mixtures

The synthetic mixtures for evaluating the accuracy of our method were created by combining reads from several single-patient sequencing samples downloaded from ENA (see Supplementary Tables S1a and S1b). For initial experiments, we have selected a subset of considered variants and pooled all mapped reads from all Fall 2020 samples belonging to these variants (Supplementary Table S1a). For experiments evaluating the accuracy at low coverages, we have subsampled these mixtures to the desired coverage. To assess the sensitivity to low abundance variants, we have created mixtures of pairs of variants, again pooling all samples belonging to a particular variant from the Fall 2020 samples. The reads corresponding to the two variants were then subsampled to achieve the desired proportions and the total average coverage of 5000. A similar procedure was used for samples representing transitions from Alpha to Delta and from Delta to Omicron. In all syntetic mixtures, the true proportions were defined as the proportion of reads originating from the samples belonging to a particular variant.

### In vitro mixture sequencing

We have created and sequenced a mixture of eight single-patient samples (Supplementary Table S2) that were previously sequenced individually at the Biomedical Research Centre of the Slovak Academy of Sciences in Bratislava, Slovakia. The samples originated from oropharyngeal swabs collected in January 2021. The sample processing and sequencing library construction were carried out as described earlier (Brejová et al, 2021), generally following the COVID-19 virus protocol (PTC 9096 v109 revF 06Feb2020; Oxford Nanopore Technologies, Oxford, UK) with some modifications. After RNA extraction for each sample, SARS-CoV-2 RNA was quantified by an RT-qPCR assay carried on QuantStudio™ 5 Real-Time PCR System (Applied Biosystem, Foster City, California, USA). Individual samples were diluted according to the obtained quantification cycle (Cq) to obtain approximately equimolar mixture (Supplementary Table S2). From this mixture of all eight samples, 11*µ*l was then used for reverse transcription. The resulting cDNA was amplified using the 2-kb primer scheme (Resende et al, 2020), and the sequencing library was constructed using a ligation kit (SQK-LSK109). A single library was used for sequencing, and thus no barcoding was performed. The library was sequenced using an R9.4.1 flow cell (FLO-MIN106) on a MinION Mk-1b device (Oxford Nanopore Technologies, Oxford, UK). Nanopore sequencing data were base called using Guppy v.4.4.1 and aligned using minimap2 v.2.13-r852 Li (2018).

### Wastewater samples

As some wastewater samples have a very uneven coverage, signal from extremely highly covered positions can overwhelm the rest of the genome. Therefore we subsample reads from such highly covered positions. In our experiments, we set coverage threshold *t* to 1000. For each read, we compute the median coverage *m*_*r*_ of the genome in the area covered by this read. The read is then chosen into the subsampled set with probability min 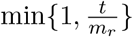. As a result, all reads are kept in regions with coverage below *t*.

To visually compare the time series obtained by our wastewater analysis with variant proportions from GISAID (as in Figure 5), we smooth the GISAID frequencies using *locally weighted linear regression* (Cleveland, 1981), as the number of samples sequenced in a single day in the location of interest can be low. The data set used for regression contains a point for each sample from a given location in GISAID. This point is (*d*, 1) for a sample from date *d* with the given variant or (*d*, 0) for any other variant. In other words, vector *X* consists of dates of the samples (converted to e.g. day difference from the beginning of year 2020), and vector *Y* contains values 0 and 1, depending on the variant of each sample. We use the Gaussian distance function 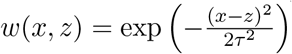, where *τ* is a smoothness parameter.

## Discussion

In this paper, we have presented a new method for estimating the proportions of SARS-CoV-2 variants in mixed samples based on a probabilistic model, which captures known genetic variability of each variant. Explicit modeling of sequencing errors and utilization of haplotype information makes the proposed approach particularly useful for current long-read sequencing technologies. We have demonstrated on synthetic mixtures that our tool gives accurate results for different sequencing technologies and with different variant combinations, including variants that were included in the background “other” profile. The proportions can be estimated even at relatively low coverage and even for variants with proportions as low as 5%. This makes it also relevant for deconvoluting coinfections in single-patient samples. Our tool can be easily adapted to other viruses where a comprehensive database of sequences belonging to individual clades is available.

Our work suggests several open problems in this area. First, we have selected the set of variants manually in this paper. This is often practical, as various authorities post lists of variants of concern. Nonetheless, automated selection of relevant variants for a given sample is an interesting problem. Some work in this direction was done by Amman et al (2022) who select variants prior to estimating their proportions.

It would be also desirable to provide some measure of confidence whether a given variant actually appears in the sample, particularly when the model predicts relatively low proportion. This can be achieved by measuring of the statistical significance of individual estimated proportions, similarly to classic regression analysis, or by requiring that reads supporting a given variant are spread along the entire genome. Highly uneven support of a variant along the genome can be caused by the presence of recombinant viruses. It would be interesting to extend our model to discover the presence of such recombinants in the sample. The ability of our model to capture long-range dependencies within reads, coupled with use of long amplicons and nanopore sequencing, is likely to contribute crucial information to detecting recombination points.

We could also extend our probabilistic model by removing various assumptions built into it. Although the entire read is in our model generated from one variant, and thus the positions in a read are not independent, once the variant is fixed, the bases in the variant profile are assumed to be independent. By building more complex variant profiles it would be possible to capture linkage between different genome positions, particularly positions within a single amplicon. Similarly, although we do not assume that the coverage is uniform at all positions, we assume that the variant proportions are the same at all positions in the genome. However, mutations occurring at primer binding sites may render some primer pairs less efficient in some variants, which violates this assumption (see e.g. changes in the ARTIC primer sets V4 and V4.1 to avoid mutations in the Delta and Omicron variants, respectively). To handle this phenomenon, our model can be extended to consider the starting position of the read as a random variable. The typical read depth profiles of individual variants necessary for this change can be estimated from single-patient samples sequenced using the same technology and primer set.

## Data Availability

All data produced in the present work are contained in the manuscript

https://github.com/fmfi-compbio/virpool

## Acknowledgements

The authors would like to thank Jozef Nosek, Rudolf Markt, Heribert Insam, and Radoslav Harman for helpful discussions.

## Funding

This research was supported by a grant from the Slovak Research and Development Agency APVV-18-0239, by grants from the Slovak grant agency VEGA 1/0463/20 (to BB), 1/0538/22 (to TV), and by the grant from the Operational Program Integrated Infrastructure ITMS:313011ATL7. The research was also supported from the European Union’s Horizon 2020 Research and Innovation Staff Exchange programme under the Marie Sklodowska-Curie grant agreement No. 872539 (PANGAIA).

## Availability of data and materials

In vitro mixture sequencing data are available from European Nucleotide Archive under project number PRJEB53383. Data from other studies are available from European Nucleotide Archive through accession numbers listed in Supplementary Table S1. We gratefully acknowledge the authors from the originating laboratories responsible for obtaining the specimens, as well as the submitting laboratories where the genome data were generated and shared via GISAID (https://www.gisaid.org/) which were used to estimate variant profiles.

## Ethics approval and consent to participate

Clinical specimens sequenced within this study were previously collected for the purpose of primary diagnosis of SARS-CoV-2 and were transferred to Biomedical Research Center of the Slovak Academy of Sciences while made unidentifiable for the researchers performing this study. The study has been approved by the Ethics\ Committee of Biomedical Research Center of the Slovak Academy of Sciences (statement no. EK/BmV-02/2020).

## Supplementary figures and tables

This table is in the attached supplementary file 1.

**Table S1:** (a) Overview of single-patient clinical samples used in synthetic mixtures. (b) Composition of synthetic mixtures and parameters of VirPool analysis. (c) Overview of wastewater samples.

**Table S2:**
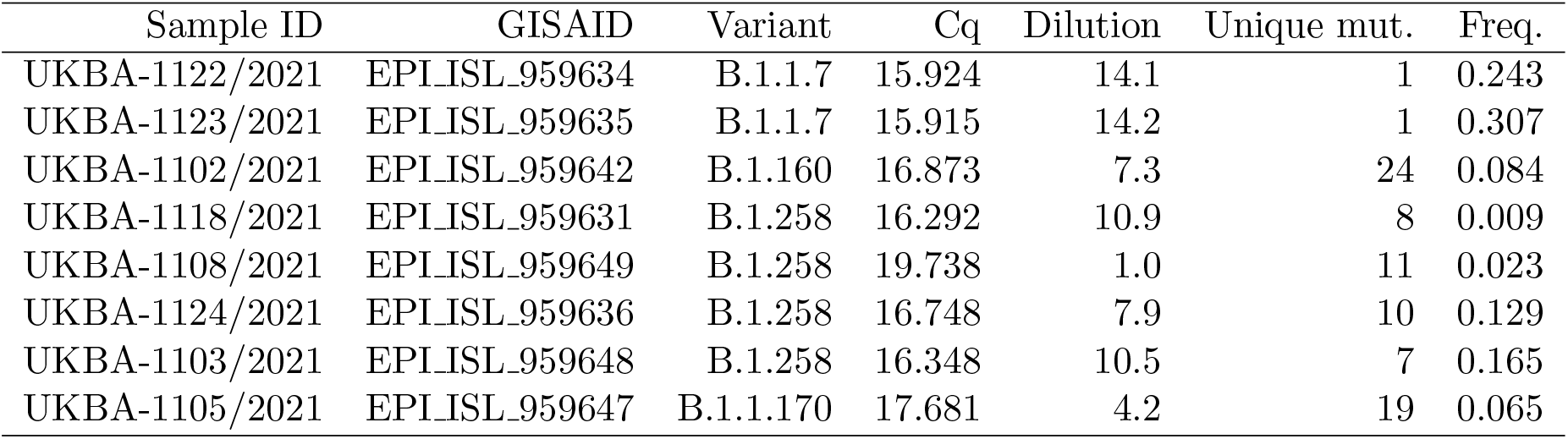
Eight single-patient samples mixed in the in vitro mixture. The dilution of each sample was computed based on the Cq value from RT-qPCR. Among mutations discovered in individual samples we have selected those that are specific to a single of these eight samples, and we report their count and the median frequency of these mutations in the mixed sample. The two B.1.1.7 samples have only a single unique mutation each, making their estimates less reliable, but their sum agrees well with proportions of mutations characteristic for this variant shown in Table S3).

**Figure S1:**
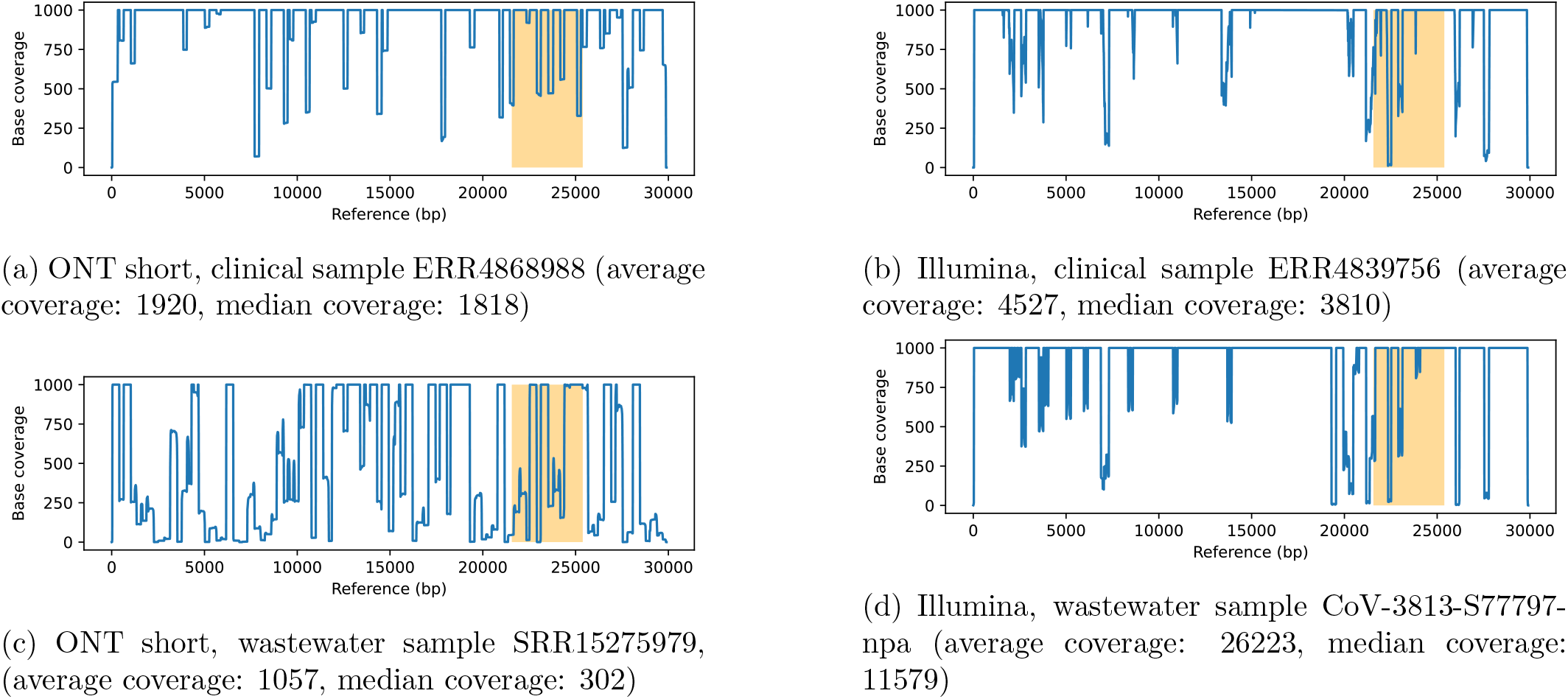
Typical coverage along the genome in clinical single-patient and wastewater samples. The yaxis is truncated at coverage of 1000. In wastewater samples, the coverage is much more variable with many regions of extremely low coverage. The Spike gene region, which contains many variant segregating mutations, is highlighted in orange.

**Table S3:** Frequencies of selected alleles in the reads from the in vitro mixed sample. We have selected all alleles which had in our variant profile probability more than 90% in B.1.1.7 and less than 1% in the other variants considered in the analysis, including the background “other” profile. We display the number and fraction of reads containing this allele and the probability in the B.1.1.7 profile. The median allele proportion is 0.524, which agrees well with the Virpool estimate of 53% for B.1.1.7. Most positions have a similar proportion of the B.1.1.7 allele, except for the last one.

| Pos | Allele | # reads | Frequency in |  |
| --- | --- | --- | --- | --- |
|  |  |  | reads | B.1.1.7 |
| 23271 | A | 12762 | 0.615 | 1.000 |
| 23709 | T | 12240 | 0.606 | 1.000 |
| 23063 | T | 12108 | 0.597 | 1.000 |
| 14676 | T | 39639 | 0.581 | 1.000 |
| 15279 | T | 38363 | 0.574 | 1.000 |
| 24506 | G | 11952 | 0.566 | 1.000 |
| 24914 | C | 11645 | 0.560 | 0.999 |
| 16176 | C | 11035 | 0.549 | 0.999 |
| 5388 | A | 24426 | 0.538 | 1.000 |
| 6954 | C | 10154 | 0.527 | 0.999 |
| 28111 | G | 31574 | 0.521 | 1.000 |
| 913 | T | 14904 | 0.502 | 1.000 |
| 3267 | T | 12638 | 0.489 | 1.000 |
| 28280 | C | 34572 | 0.487 | 0.998 |
| 27972 | T | 27369 | 0.486 | 1.000 |
| 28977 | T | 26171 | 0.486 | 1.000 |
| 28048 | T | 28749 | 0.484 | 0.998 |
| 28282 | A | 33981 | 0.477 | 0.998 |
| 28281 | T | 33260 | 0.445 | 0.998 |
| 11296 | G | 456 | 0.053 | 0.918 |

**Table S4:**
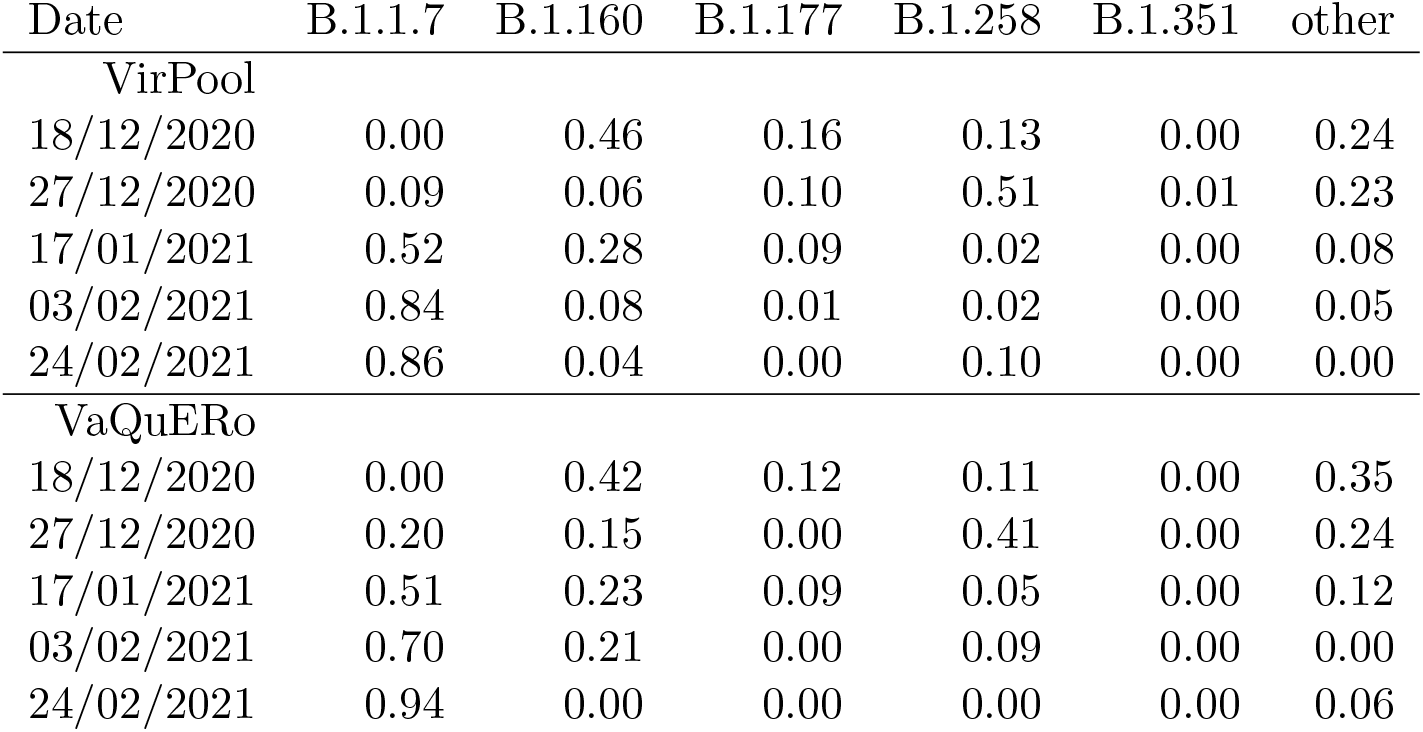
The estimated proportions in wastewater samples from a plant near Bischofshofen, Austria (federal state of Salzburg) (Amman et al, 2022). Comparison of predictions with VirPool and VaQuERo (Amman et al, 2022). Column “other” for VaQuERo includes lineages B.1.221, B.1.1.232, and B.1.1.153 as well as other unidentified lineages.

**Table S5:**
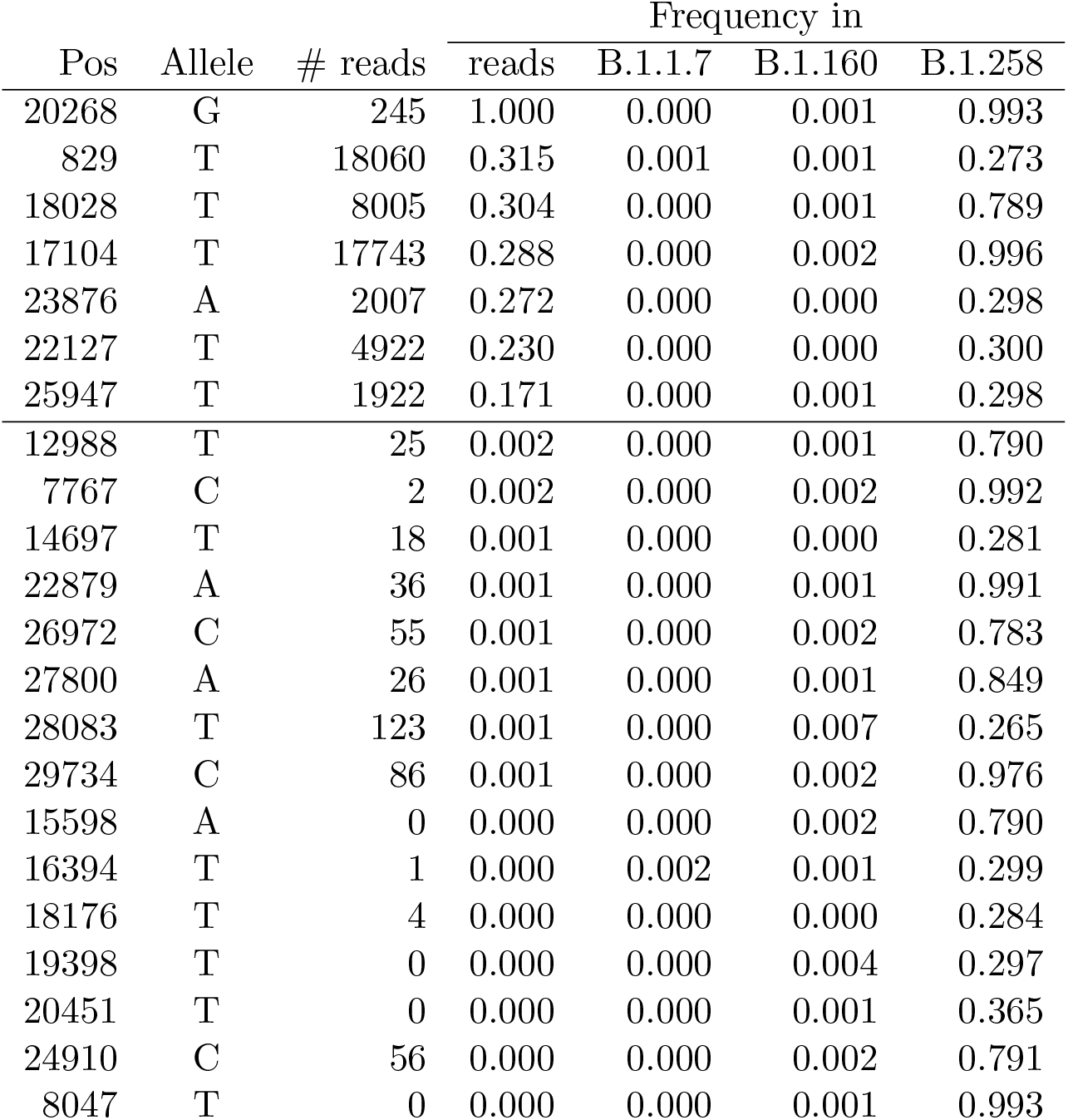
Frequencies of selected alleles in the Austrian wastewater sample from February 24, 2021. We have selected all alleles which had in our variant profile probability less that 1% for both B.1.1.7 and B.1.160 and probability at least 25% in B.1.258. We also display the number and fraction of reads containing this allele and the frequencies from these profiles. The first seven rows support a high percentage of B.1.258 variant.

**Table S6:**
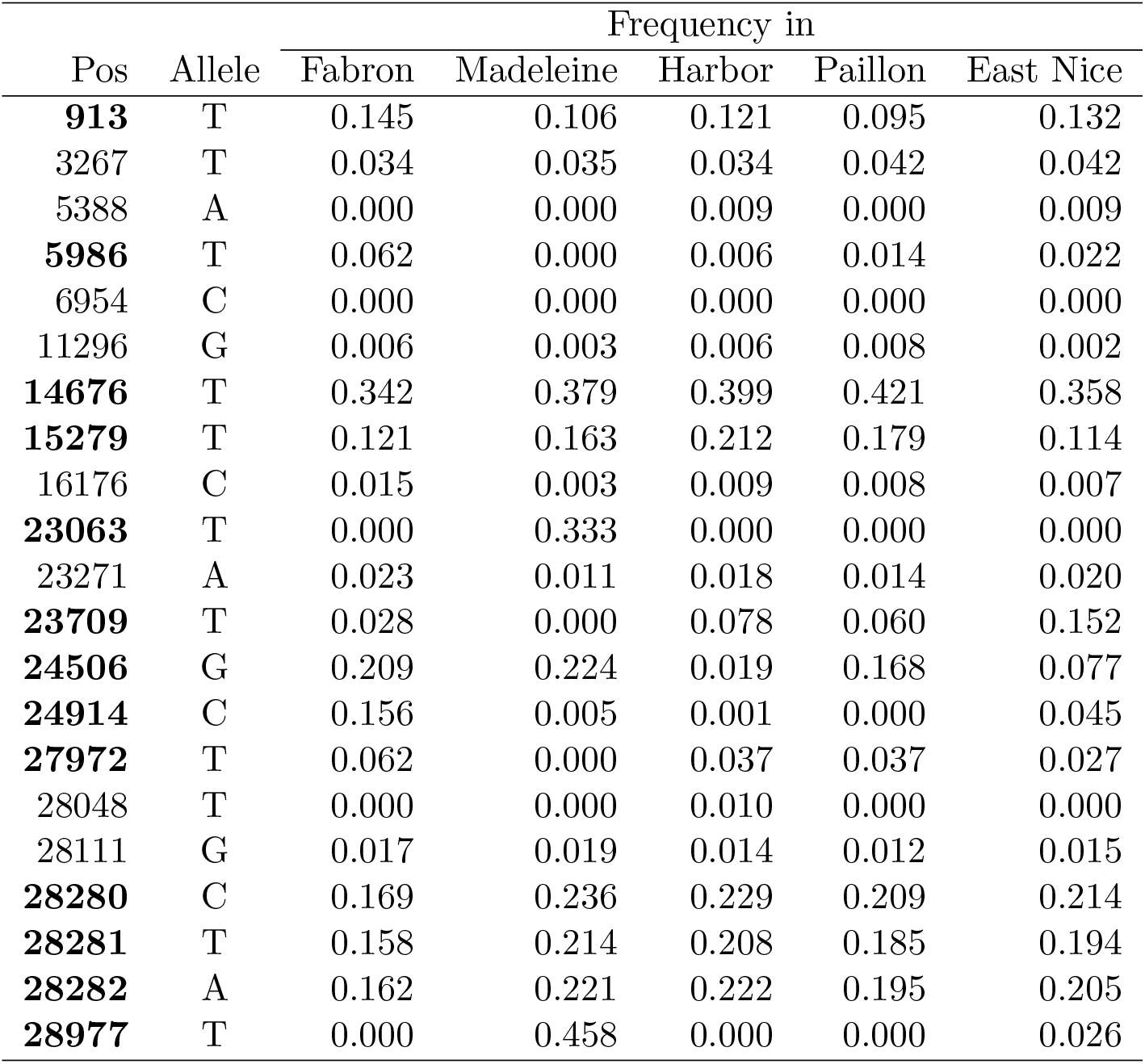
Frequencies of selected alleles in the Nice wastewater samples from October 2020. We have selected all alleles which had in our variant profile probability less that 1% for both B.1.160 and “other” and probability at least 90% in B.1.1.7. We display the fraction of reads containing each allele in the samples from the five locations where VirPool estimated at least 7% of the Alpha variant. Positions where at least one of the five samples have allele proportion at least 5% are highlighted in boldface.

**Table S7:**
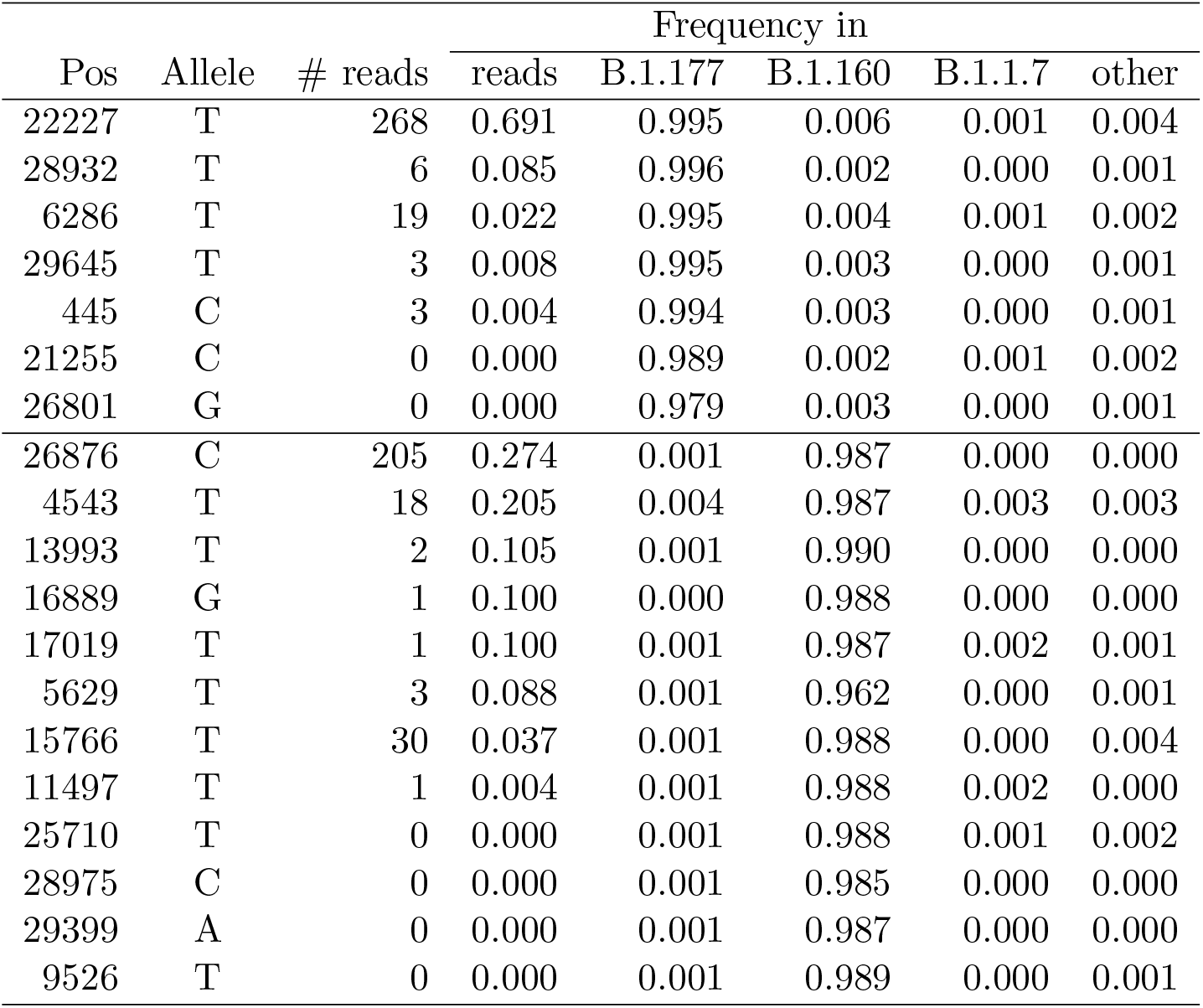
Frequencies of selected alleles in the Nice wastewater samples from March 2021, Harbor location. The top part of the table contains all alleles which had in our variant profile probability less that 1% for B.1.160, B.1.1.7 and “other” and probability at least 90% in B.1.177. Similarly, the bottom part contains all alleles with probability at least 90% in B.1.160 and less than 1% in B.1.177, B.1.1.7 and “other”. We display the count and fraction of reads containing each allele in this sample as well as allele probabilities in these profiles.

## Notes

### Competing Interest Statement

The authors have declared no competing interest.

### Author Declarations

Ethics Committee of Biomedical Research Center of the Slovak Academy of Sciences gave ethical approval for this work (statement no. EK/BmV-02/2020)

## References

1 Agrawal S, Orschler L, Lackner S (2021) Metatranscriptomic Analysis Reveals SARS-CoV-2 Mutations in Wastewater of the Frankfurt Metropolitan Area in Southern Germany. Microbiology Resource Announcements 10(15):e00280–21

2 Agrawal S, Orschler L, Schubert S, Zachmann K, Heijnen L, Tavazzi S, Gawlik BM, de Graaf M, Medema G, Lackner S (2022) Prevalence and circulation patterns of SARS-CoV-2 variants in European sewage mirror clinical data of 54 European cities. Water Research 214:118162

3 Ahn S, Vikalo H (2018) aBayesQR: A Bayesian Method for Reconstruction of Viral Populations Characterized by Low Diversity. Journal of Computational Biology 25(7):637–648

4 Amman F, Markt R, Endler L, Hupfauf S, Agerer B, Schedl A, Richter L, Zechmeister M, Bicher M, Heiler G, et al (2022) National-scale surveillance of emerging SARS-CoV-2 variants in wastewater. medRxiv 2022.01.14.21267633

5 Bibby K, Bivins A, Wu Z, North D (2021) Making waves: Plausible lead time for wastewater based epidemiology as an early warning system for COVID-19. Water Research 202:117438

6 Brejová B, Boršová K, Hodorová V, Čabanová V, Gafurov A, Fričová D, Neboháčová M, Vinař T, Klempa B, Nosek J (2021) Nanopore sequencing of sars-cov-2: Comparison of short and long pcr-tiling amplicon protocols. PloS One 16(10):e0259277

7 Bridle JS (1990) Probabilistic interpretation of feedforward classification network outputs, with relationships to statistical pattern recognition. In: Neurocomputing, Springer, pp 227–236

8 Brunner FS, Brown MR, Bassano I, Denise H, Khalifa MS, Wade M, Kevill JL, Jones DL, Farkas K, Jeffries AR, et al (2022) City-wide wastewater genomic surveillance through the successive emergence of SARS-CoV-2 Alpha and Delta variants. medRxiv 2022.02.16.22269810

9 Cleveland WS (1981) LOWESS: A program for smoothing scatterplots by robust locally weighted regression. American Statistician 35(1):54

10 Crits-Christoph A, Kantor RS, Olm MR, Whitney ON, Al-Shayeb B, Lou YC, Flamholz A, Kennedy LC, Greenwald H, Hinkle A, Hetzel J, Spitzer S, Koble J, Tan A, Hyde F, Schroth G, Kuersten S, Banfield JF, Nelson KL (2021) Genome sequencing of sewage detects regionally prevalent SARS-CoV-2 variants. mBio 12(1):e02703–20

11 De Maio N, Walker C, Borges R, Weilguny L, Slodkowicz G, Goldman N (2020) Issues with SARS-CoV-2 sequencing data. virologicalorg https://virological.org/t/issues-with-sars-cov-2-sequencing-data/473/1. Accessed 13 June 2022

12 Eden JS, Sim E (2020) SARS-CoV-2 genome sequencing using long pooled amplicons on Illumina platforms. protocolsio DOI 10.17504/protocols.io.befyjbpw

13 Elbe S, Buckland-Merrett G (2017) Data, disease and diplomacy: GISAID’s innovative contribution to global health. Global Challenges 1(1):33–46

14 Ellmen I, Lynch MD, Nash D, Cheng J, Nissimov JI, Charles TC (2021) Alcov: Estimating variant of concern abundance from SARS-CoV-2 wastewater sequencing data. medRxiv 2021.06.03.21258306

15 Eriksson N, Pachter L, Mitsuya Y, Rhee SY, Wang C, Gharizadeh B, Ronaghi M, Shafer RW, Beerenwinkel N (2008) Viral population estimation using pyrosequencing. PLoS Computational Biology 4(5):e1000074

16 Fontenele RS, Kraberger S, Hadfield J, Driver EM, Bowes D, Holland LA, Faleye TO, Adhikari S, Kumar R, Inchausti R, et al (2021) High-throughput sequencing of SARS-CoV-2 in wastewater provides insights into circulating variants. medRxiv 2021.01.22.21250320

17 Freed NE, Vlkova M, Faisal MB, Silander OK (2020) Rapid and inexpensive whole-genome sequencing of SARS-CoV-2 using 1200 bp tiled amplicons and Oxford Nanopore Rapid Barcoding. Biology Methods&Protocols 5(1):bpaa014

18 Gafurov A, Baláž A, Vinař T, Brejová B (2021) Estimation of proportions of SARS-CoV-2 variants in a mixed sequencing sample. CEUR Workshop Proceedings 2962:301–307

19 Hillary LS, Farkas K, Maher KH, Lucaci A, Thorpe J, Distaso MA, Gaze WH, Paterson S, Burke T, Connor TR, McDonald JE, Malham SK, Jones DL (2021) Monitoring SARS-CoV-2 in municipal wastewater to evaluate the success of lockdown measures for controlling COVID-19 in the UK. Water Research 200:117214

20 Hrudey SE, Conant B (2022) The devil is in the details: emerging insights on the relevance of wastewater surveillance for SARS-CoV-2 to public health. Journal of Water and Health 20(1):246–270

21 Izquierdo-Lara R, Elsinga G, Heijnen L, Oude Munnink BB, Schapendonk CM, Nieuwenhuijse D, Kon M, Lu L, Aarestrup FM, Lycett S, Medema G, Koopmans MP, De Graaf M (2021) Monitoring SARS-CoV-2 circulation and diversity through community wastewater sequencing, the netherlands and belgium. Emerging Infectious Diseases 27(5):1405–1415

22 Jahn K, Dreifuss D, Topolsky I, Kull A, Ganesanandamoorthy P, Fernandez-Cassi X, Bänziger C, Stachler E, Fuhrmann L, Philipp Jablonski K, Chen C, Aquino C, Stadler T, Ort C, Kohn T, Julian TR, Beerenwinkel N (2021) Detection of SARS-CoV-2 variants in Switzerland by genomic analysis of wastewater samples. medRxiv 2021.01.08.21249379

23 La Rosa G, Mancini P, Bonanno Ferraro G, Veneri C, Iaconelli M, Lucentini L, Bonadonna L, Brusaferro S, Brandtner D, Fasanella A, Pace L, Parisi A, Galante D, Suffredini E (2021) Rapid screening for SARS-CoV-2 variants of concern in clinical and environmental samples using nested RT-PCR assays targeting key mutations of the spike protein. Water Research 197:117104

24 Li H (2018) Minimap2: pairwise alignment for nucleotide sequences. Bioinformatics 34(18):3094–3100

25 Loman N, et al (2020) ARTIC nanopore protocol for nCoV2019 novel coronavirus. https://github.com/artic-network/artic-ncov2019. Accessed 13 June 2022.

26 Nemudryi A, Nemudraia A, Wiegand T, Surya K, Buyukyoruk M, Cicha C, Vanderwood KK, Wilkinson R, Wiedenheft B (2020) Temporal detection and phylogenetic assessment of SARS-CoV-2 in municipal wastewater. Cell Reports Medicine 1(6):100098

27 Pechlivanis N, Tsagiopoulou M, Maniou MC, Togkousidis A, Mouchtaropoulou E, Chassalevris T, Chaintoutis SC, Petala M, Kostoglou M, Karapantsios T, et al (2022) Detecting SARS-CoV-2 lineages and mutational load in municipal wastewater and a use-case in the metropolitan area of Thessaloniki, Greece. Scientific Reports 12(1):2659

28 Quick J, Grubaugh ND, Pullan ST, Claro IM, Smith AD, Gangavarapu K, Oliveira G, Robles-Sikisaka R, Rogers TF, Beutler NA, Burton DR, Lewis-Ximenez LL, de Jesus JG, Giovanetti M, Hill SC, Black A, Bedford T, Carroll MW, Nunes M, Alcantara LC, Sabino EC, Baylis SA, Faria NR, Loose M, Simpson JT, Pybus OG, Andersen KG, Loman NJ (2017) Multiplex PCR method for MinION and Illumina sequencing of Zika and other virus genomes directly from clinical samples. Nature Protocols 12(6):1261–1276

29 Rambaut A, Holmes EC, O’Toole Á, Hill V, McCrone JT, Ruis C, du Plessis L, Pybus OG (2020) A dynamic nomenclature proposal for SARS-CoV-2 lineages to assist genomic epidemiology. Nature Microbiology 5(11):1403–1407

30 Resende PC, Motta FC, Roy S, Appolinario L, Fabri A, Xavier J, Harris K, Matos AR, Caetano B, Orgeswalska M, et al (2020) SARS-CoV-2 genomes recovered by long amplicon tiling multiplex approach using nanopore sequencing and applicable to other sequencing platforms. bioRxiv 2020.04.30.069039

31 Rios G, Lacoux C, Leclercq V, Diamant A, Lebrigand K, Lazuka A, Soyeux E, Lacroix S, Fassy J, Couesnon A, et al (2021a) Characteristic mutations of SARS-CoV-2 variants. https://github.com/ucagenomix/cagablea/blob/main/data_base/agg_data_lineage_All_2019-12-15-2020-12-31_top_2000.csv. Accessed 13 June 2022.

32 Rios G, Lacoux C, Leclercq V, Diamant A, Lebrigand K, Lazuka A, Soyeux E, Lacroix S, Fassy J, Couesnon A, et al (2021b) Monitoring SARS-CoV-2 variants alterations in Nice neighborhoods by wastewater nanopore sequencing. The Lancet Regional Health-Europe 10:100202

33 Safford HR, Shapiro K, Bischel HN (2022) Wastewater analysis can be a powerful public health tool—if it’s done sensibly. Proceedings of the National Academy of Sciences 119(6):e2119600119

34 Van Poelvoorde LAE, Delcourt T, Coucke W, Herman P, De Keersmaecker SCJ, Saelens X, Roosens NHC, Vanneste K (2021) Strategy and performance evaluation of low-frequency variant calling for SARS-CoV-2 using targeted deep Illumina sequencing. Frontiers in Microbiology 12:747458

35 Virtanen P, Gommers R, Oliphant TE, Haberland M, Reddy T, Cournapeau D, Burovski E, Peterson P, Weckesser W, Bright J, van der Walt SJ, Brett M, Wilson J, Millman KJ, Mayorov N, Nelson ARJ, Jones E, Kern R, Larson E, Carey CJ, Polat I, Feng Y, Moore EW, VanderPlas J, Laxalde D, Perktold J, Cimrman R, Henriksen I, Quintero EA, Harris CR, Archibald AM, Ribeiro AH, Pedregosa F, van Mulbregt P, SciPy 10 Contributors (2020) SciPy 1.0: Fundamental algorithms for scientific computing in python. Nature Methods 17:261–272

36 Wurtz N, Revol O, Jardot P, Giraud-Gatineau A, Houhamdi L, Soumagnac C, Annessi A, Lacoste A, Colson P, Aherfi S, La Scola B (2021) Monitoring the circulation of SARS-CoV-2 variants by genomic analysis of wastewater in Marseille, south-east France. Pathogens 10(8):1042

37 Xie Y, Challis JK, Oloye FF, Asadi M, Cantin J, Brinkmann M, McPhedran KN, Hogan N, Sadowski M, Jones PD, Landgraff C, Mangat C, Servos MR, Giesy JP (2022) RNA in municipal wastewater reveals magnitudes of COVID-19 outbreaks across four waves driven by SARS-CoV-2 variants of concern. ACS ES&T Water

38 Zagordi O, Bhattacharya A, Eriksson N, Beerenwinkel N (2011) ShoRAH: estimating the genetic diversity of a mixed sample from next-generation sequencing data. BMC Bioinformatics 12:119

39 Zhu C, Byrd RH, Lu P, Nocedal J (1997) Algorithm 778: L-BFGS-B: Fortran Subroutines for Large-Scale Bound-Constrained Optimization. ACM Transactions on Mathematical Software 23(4):550–560

